# Mining Social Media Data for Influenza Vaccine Effectiveness Using a Large Language Model and Chain-of-Thought Prompting

**DOI:** 10.1101/2025.03.26.25324701

**Authors:** Dongfang Xu, Guillermo López García, Karen O’Connor, Haily Holston, Ari Z Klein, Ivan Flores Amaro, Matthew Scotch, Graciela Gonzalez-Hernandez

## Abstract

Influenza vaccine effectiveness (VE) estimation plays a critical role in public health decision-making by quantifying the real-world impact of vaccination campaigns and guiding policy adjustments. Current approaches to VE estimation are constrained by limited population representation, selection bias, and delayed reporting. To address some of these gaps, we propose leveraging large language models (LLMs) with few-shot chain-of-thought (CoT) prompting to mine social media data for real-time influenza VE estimation. We annotated over 4,000 tweets from the 2020–2021 flu season using structured guidelines, achieving high inter-annotator agreement. Our best prompting strategy achieves F_1_ scores above 87% for identifying influenza vaccination status and test outcomes, outperforming traditional supervised fine-tuning methods by large margins. These findings indicate that LLM-based prompting approaches effectively identify relevant social media information for influenza VE estimation, offering a valuable real-time surveillance tool that complements traditional epidemiological methods.

## INTRODUCTION

### Background and Significance

The 2024–2025 influenza season has been classified as the most severe season since 2017–2018 [1]. According to the U.S. Centers for Disease Control and Prevention (CDC), the overall burden of influenza for the 2024-2025 season includes at minimum, 41 million flu illnesses, 19 million flu-related medical visits, 540,000 hospitalizations, and 23,000 deaths since October 1, 2024 [2]. The primary public health intervention against seasonal influenza is the annual flu vaccine, which is developed and manufactured several months before the flu season. However, flu vaccine effectiveness (VE) varies annually and even within the same season, emphasizing the need for real-time evaluation of its impact. Accurate VE estimates are vital for assessing vaccination program outcomes, enabling public health authorities to estimate influenza-related morbidity and mortality accurately, and informing targeted control strategies and outreach activities, especially during seasons with reduced vaccine effectiveness [3].

Traditionally, the CDC estimates flu VE annually through the US Flu VE Network [5]. This network operates only at select clinics within a limited number of states, enrolling approximately 1,000 participants per site each year with influenza-like illness using a test-negative design (see Figure 1). The CDC provides VE estimates at the end of each flu season, with interim estimates published by late February or thereafter. Although regarded as the gold standard in the U.S., the CDC estimation have two notable limitations: (1) limited geographic and participant representation, and (2) delayed publication of interim reports during the flu season. In this study, we propose leveraging natural language processing (NLP) to analyze social media data as a complementary approach to address these limitations. We hypothesize that social media data, abundantly available nationwide and in near real-time, can enhance VE estimation by expanding geographic coverage and timeliness. Prior studies have demonstrated the effectiveness of NLP techniques applied to social media data for epidemiological research [6, 7, 8]. To our knowledge, this is the first study to propose an automated, near-real-time approach to estimating flu VE using patient reports posted publicly on social media.

**Figure 1:**
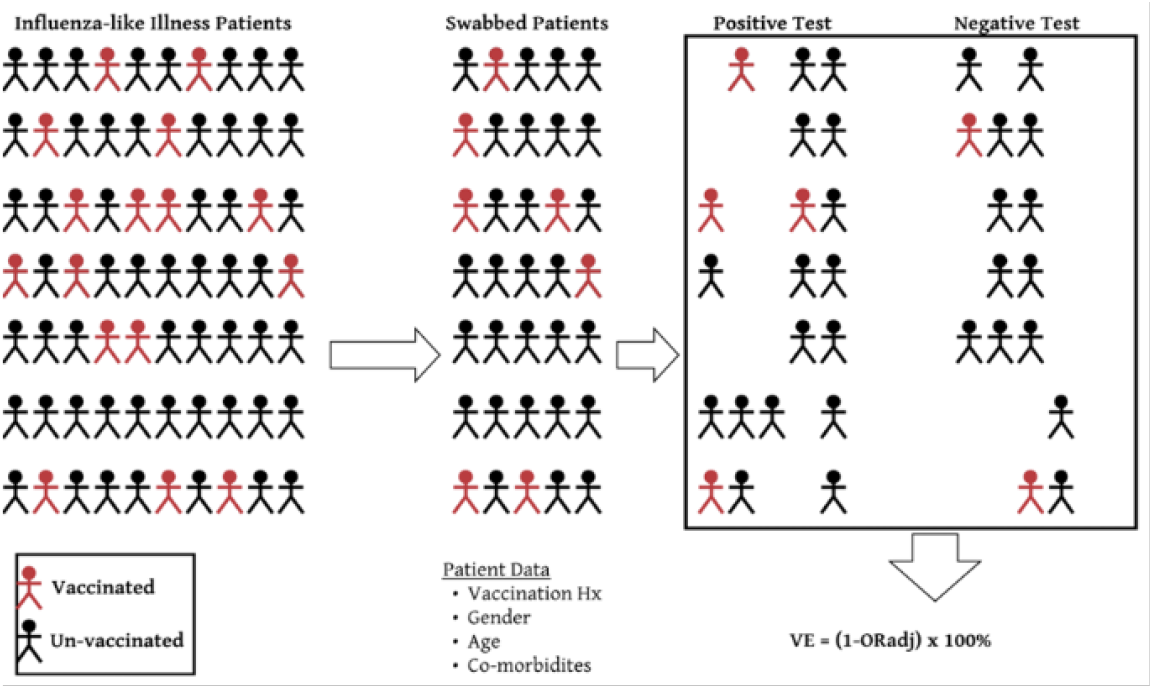
Test negative design for influenza vaccine effectiveness. Diagram modified from [4]. Abbreviations: Hx= history; VE = vaccine effectiveness

To mimic the “test-negative design” used by the CDC [4], we first identify users that have publicly posted reports on X (formerly known as Twitter) of having been tested for flu (referred to as “Swabbed Patients” in Figure 1), as well as their test results. Further, from all their publicly available posts, we extract whether they have been vaccinated or not. We place each user into one of four possible flu-test-and-vaccine categories, essential for calculating the odds ratio (OR) as defined by equation (1): unvaccinated and flu-negative (*Unvac-Neg*), unvaccinated and flu-positive (*Unvac-Pos*), vaccinated and flu-negative (*Vac-Neg*), and vaccinated and flu-positive (*Vac-Pos*). We then calculate: *VE* = (1 − *OR*) *×* 100%. This *OR* can be further adjusted based on patient characteristics.

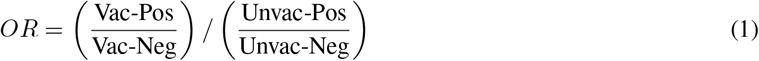

## OBJECTIVE

The overall objective of this study is to identify influenza test outcomes and vaccination status from social media data for real-time estimation of flu VE. The focus of this paper is to detail the development and validation of the NLP approaches we develop as a first step towards the overall objective. First, we established a structured annotation guideline and annotated over 4,000 tweets from a recent influenza season. Building on this guideline and annotation, we then explored a LLM based approach with various prompting strategies and compared them against traditional supervised fine-tuning methods.

## MATERIALS and METHODS

### Data collection

We collected tweets from X and identified a pool of users of interest from the 2019–2020 and 2020–2021 flu seasons. We first used keywords against the X API such as *influenza* and *flu* to retrieve an initial collection of tweets. To enhance precision and reduce irrelevant results, we subsequently developed targeted regular expressions to capture more specific phrases indicative of flu vaccination status and testing outcome, including expressions like *had a positive flu test* or *received the flu vaccine*. Ultimately, we collected 432,278 tweets related to flu vaccination and 64,166 tweets related to flu testing. Recognizing that our system is designed for the post-COVID-19 context, where tweets often reference both flu and COVID-19 tests, we specifically sampled and annotated 4,216 tweets from the 2020–2021 flu season for better distinguishing influenza-related from COVID-19-related events. We reserved the entire collection from the 2019-2020 flu season for validation.

### Data Annotation

To ensure high-quality data for developing our system, we established a structured annotation guideline for identifying flu vaccine and flu test reports in tweets. This guideline carefully addresses nuanced expressions related to vaccination and flu testing behaviors, distinguishes between direct personal experiences and general references, and differentiates flu-related events according to their temporal context—current flu season, past flu season, or anticipated in the future. Recognizing that the flu season can extend beyond standard calendar dates, we defined a fixed temporal window, from September 1 of the start year to August 31 of the following year, to maintain consistency.

Through a linguistic analysis of approximately 100 tweets, we identified distinct patterns differentiating flu vaccine tweets from flu test tweets. Flu vaccine discussions were typically explicit, clearly indicating the user’s vaccination status or experiences in the current season. In contrast, flu test references tended to be indirect or contextual, often marked by uncertainty and diagnostic ambiguity. Moreover, tweets mentioning flu tests commonly referenced past experiences rather than explicitly stating a status relevant to the current flu season. This finding underscores the necessity of precisely interpreting temporal context during our annotation process.

### Flu Vaccine Annotation

We developed a taxonomy that categorizes tweets based on the user’s self-reported vaccination status: *Currently-Vaccinated*: The user explicitly states receiving a flu shot in the current flu season.

#### Currently-Unvaccinated

The user explicitly states that they have not received or plan not to receive a flu shot. Statements reflecting past behaviors are included if they imply a continuous pattern and allow inference about the user’s current vaccination status.

#### Previously-Vaccinated

The user mentions receiving a flu shot during a previous flu season without indicating any recent vaccination updates.

#### Possibly-Vaccinated

The user indicates intentions or considerations to receive a flu shot but provides no clear evidence of actual vaccination.

#### Other

The tweet references flu vaccination without specifying the user’s personal vaccination status, including mentions of other individuals’ vaccination statuses (such as their child), advocacy for flu shots, or general vaccine-related information.

### Flu Test Annotation

We developed a similar taxonomy for annotating flu test outcomes. Notably, past flu test mentions predominantly reflect negative results (approximately 80%), unlike past vaccination mentions, which primarily indicate positive status (received vaccination). To accurately represent this distinction, we categorized past flu test outcomes separately into *Positive* and *Negative*. Similar to flu vaccine, we also annotated *Possibly-Tested*, but only found less than 1% of tweets with this label, so we merged it to *Other* label. Specifically, we have the following labels:

#### Currently-Positive

The user explicitly reports a recent positive flu test result or diagnosis.

#### Currently-Negative

The user explicitly reports a recent negative flu test result or a diagnosis excluding the flu.

#### Previously-Positive

The user mentions testing positive for flu or receiving a flu diagnosis in a previous season, with no indication of their current flu test status.

#### Previously-Negative

The user mentions testing negative or not being diagnosed with flu in a previous season, without referencing their current flu test status.

#### Other

The tweet references flu testing without specifying the user’s personal test status, including references to another individual’s test results, encouragement for others to get tested, general discussions about flu tests, or ambiguous statements where the user’s test status remains unclear.

Two annotators, both with biomedical backgrounds and experience in social media research, each independently annotated the same 300 tweets for flu testing and the same 300 tweets for flu vaccination status. They identified and discussed any annotating discrepancies to reach a consensus label. Before resolution, inter-annotator agreement (IAA), measured using the *F*_1_ score, was *F*_1_ = 0.91 for flu test labels and *F*_1_ = 0.82. Table 1 shows the statistics of flu vaccine and flu test labels in our annotations. Because the computation of odds ratio requires only current-season flu vaccination and testing, we also explicitly calculated IAA for *Currently-Vaccinated, Currently-Unvaccinated, Currently-Positive*, and *Currently-Negative*. In this subset, the IAA scores were *F*_1_ = 0.86 for flu tests and *F*_1_ = 0.84 for flu vaccinations. All these IAA scores indicate a moderate-to-strong level of agreement [9]. Notably, annotations for *Previously-Vaccinated* and *Possibly-Vaccinated* labels showed greater ambiguity with other labels, resulting in a lower overall IAA score for the complete flu vaccination annotations compared to current-season annotations. After resolving disagreements, the annotators independently annotated the remaining 3,616 tweets.

**Table 1:** Distribution of annotated labels for flu vaccine and flu test outcome in tweets collected from the 2020–2021 influenza season. We show counts and proportions (%) of each label category.

| Flu Vaccine |  |  | Flu Test |  |  |
| --- | --- | --- | --- | --- | --- |
| Label | Counts | Prop (%) | Label | Counts | Prop (%) |
| Currently-Vaccinated | 581 | 20.68% | Currently-Positive | 87 | 6.18% |
| Currently-Unvaccinated | 709 | 25.24% | Currently-Negative | 116 | 8.24% |
| Previously-Vaccinated | 186 | 6.62% | Previously-Positive | 41 | 2.91% |
| Possibly-Vaccinated | 324 | 11.53% | Previously-Negative | 161 | 11.44% |
| Other | 1009 | 35.92% | Other | 1002 | 71.22% |

### Approach

We frame the identification of flu vaccination and flu test reports as two separate multiclass classification tasks. Given a tweet, we aim to assign one of the labels described in the Data Annotation section. Recent advancements in text classification have shifted from supervised fine-tuning methods toward in-context learning (ICL) approaches using LLMs [10]. ICL allows LLMs to perform novel NLP tasks without updating model parameters, relying instead on task-specific instructions and a few input-output examples (also known as “few-shot prompting”). For complex reasoning tasks, such as commonsense reasoning, chain-of-thought (CoT) prompting [11] requires LLMs to break down problems into sequential, intermediate reasoning steps before reaching a final conclusion rather than directly mapping inputs to outputs. Both ICL and CoT prompting methods have demonstrated strong performance, achieving promising or state-of-the-art results across various NLP tasks.

In this study, we utilized the LLaMA-3-70B-Instruct model together with an ICL approach combined with CoT prompting to automatically identify flu vaccination status and flu test reports from tweets.

### In Context Learning with Chain-of Thought Prompting

To design prompts for our tasks, we adopted the standard ICL framework [12] and the CoT prompting strategy [11]. Each complete prompt consisted of task instructions, several example tweets paired with their respective classification labels and reasoning steps, and the target tweet to be classified. The task instructions clearly defined the classification problem, specified the input format (tweet content and posting date), detailed the output labels, and outlined the step-by-step reasoning for identifying flu-related events. To ensure consistency in classification, we explicitly specified the output format. For the few-shot examples, we randomly selected two tweets per label, aiming to cover a variety of flu-related linguistic patterns and reasoning paths. Each example included the tweet content, the posting date, a detailed sequence of reasoning steps that describe the decision-making process, and the associated classification label. The inclusion of reasoning steps aimed to guide the model toward structured analytical reasoning rather than relying solely on superficial keyword detection.

For flu vaccine status, we designed prompts to directly predict one of the five distinct flu vaccine labels. An example of this prompting approach is provided in Table 2. Identifying flu test outcomes required a more nuanced approach due to the complexity of the task, specifically the need to distinguish both the test outcome and the relevant flu season. To address this, we decomposed the task into two sequential steps. In the first step, we prompted the LLM to identify the flu test outcome (positive, negative, or other) without considering temporal information. In the second step, the LLM was prompted to determine whether the identified test occurred during the current flu season (yes, no, or other), based on temporal indicators within the tweet and its posting date. Both steps utilized similar prompting strategies, with adjustments made to task instructions, reasoning guidance, and output labels to match the specific goals of each step. Ultimately, the outputs from these two steps were integrated to ascertain the final flu test classification. An example of this two-step prompting strategy is illustrated in Table 3.

**Table 2:** Example of ICL with CoT prompting for flu vaccine classification. In the **Instruction** row, bolded text indicates reasoning instructions, while “… “ represents placeholders for labels and their definitions provided in the flu vaccine annotation section. **Input, Reasoning**, and **Label** represent the prompts for one example.

|  |  |
| --- | --- |
| **Instruction** | Given a tweet and its posting date between September 2020 and August 2021, determine the user’s flu vaccine status for the current flu season. **Follow these steps: first, identify any flu vaccine–related texts and temporal expressions, then interpret these references in the context of the tweet and posting date.** Return exactly one of the following five labels, defined as follows: ...<br>Output Format:<br><label> |
| **Input** | *Posting Date:* 2020-12-14<br>*Tweet:* Getting a flu shot before working out for the day is a bad idea<br>Let’s think this step by step: |
| **Reasoning** | The user directly states they got a flu shot on the same day “before working out for the day”. This confirms they have received the flu vaccine for the current season. |
| **Label** | **Currently-Positive** |
| ... | ... |
| **Query** | *Posting Date:* 2021-01-05<br>*Tweet:* I got the flu shot last Oct for my Sister<br>Let’s think this step by step: |

**Table 3:**
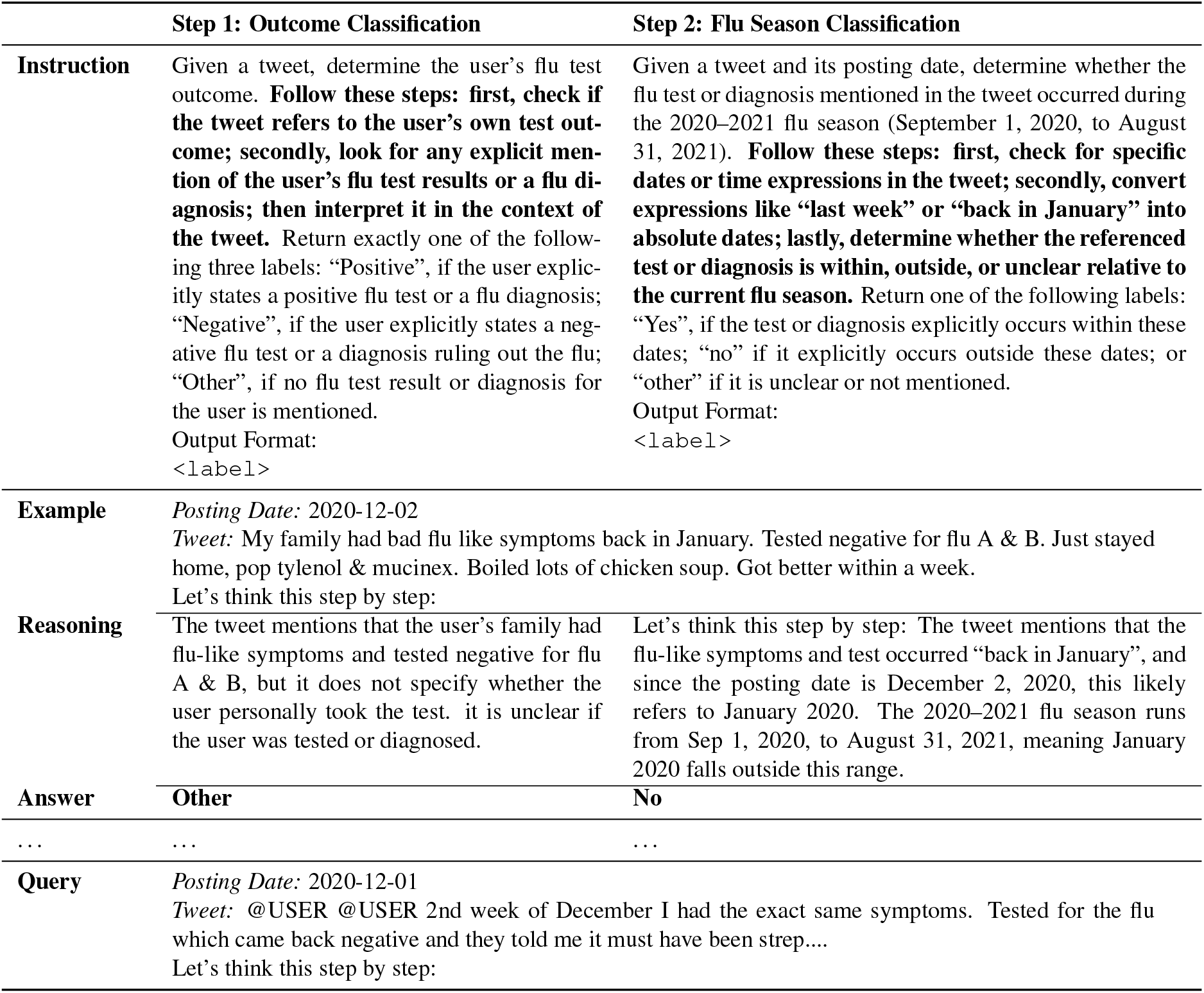
Two-step prompting for **flu test** classification. Bold in the **Instruction** row indicates reasoning instructions.

|  | Step 1: Outcome Classification | Step 2: Flu Season Classification |
| --- | --- | --- |
| **Instruction** | Given a tweet, determine the user’s flu test outcome. **Follow these steps: first, check if the tweet refers to the user’s own test outcome; secondly, look for any explicit mention of the user’s flu test results or a flu diagnosis; then interpret it in the context of the tweet.** Return exactly one of the following three labels: “Positive”, if the user explicitly states a positive flu test or a flu diagnosis; “Negative”, if the user explicitly states a negative flu test or a diagnosis ruling out the flu; “Other”, if no flu test result or diagnosis for the user is mentioned.<br>Output Format:<br><label> | Given a tweet and its posting date, determine whether the flu test or diagnosis mentioned in the tweet occurred during the 2020–2021 flu season (September 1, 2020, to August 31, 2021). **Follow these steps: first, check for specific dates or time expressions in the tweet; secondly, convert expressions like “last week” or “back in January” into absolute dates; lastly, determine whether the referenced test or diagnosis is within, outside, or unclear relative to the current flu season.** Return one of the following labels: “Yes”, if the test or diagnosis explicitly occurs within these dates; “no” if it explicitly occurs outside these dates; or “other” if it is unclear or not mentioned.<br>Output Format:<br><label> |
| **Example** | *Posting Date:* 2020-12-02<br>*Tweet:* My family had bad flu like symptoms back in January. Tested negative for flu A & B. Just stayed home, pop tylenol & mucinex. Boiled lots of chicken soup. Got better within a week.<br>Let’s think this step by step: |  |
| **Reasoning** | The tweet mentions that the user’s family had flu-like symptoms and tested negative for flu A & B, but it does not specify whether the user personally took the test. it is unclear if the user was tested or diagnosed. | Let’s think this step by step: The tweet mentions that the flu-like symptoms and test occurred “back in January”, and since the posting date is December 2, 2020, this likely refers to January 2020. The 2020–2021 flu season runs from Sep 1, 2020, to August 31, 2021, meaning January 2020 falls outside this range. |
| **Answer** | **Other** | **No** |
| ... | ... | ... |
| **Query** | *Posting Date:* 2020-12-01<br>*Tweet:* @USER @USER 2nd week of December I had the exact same symptoms. Tested for the flu which came back negative and they told me it must have been strep....<br>Let’s think this step by step: |  |

### Baseline

To evaluate the effectiveness of the CoT few-shot prompting method, we compared it against a traditional supervised baseline using Bidirectional Encoder Representations from Transformers (BERT) [13]. BERT is a smaller-scale language model specifically designed for natural language understanding tasks. In contrast to autoregressive models such as GPT-3 [10] and LLaMA [14], BERT is pretrained on relatively smaller datasets using a masked language modeling objective. While large language models (LLMs) provide broader capabilities at greater computational costs, BERT is more efficient for fine-tuning task-specific NLP tasks and was widely utilized before the recent rise of LLMs. In this study, we applied BERT within a text classification framework and explored three variants of BERT-based models:

#### BERT-large

The large-scale version of BERT released in [13], comprising approximately 340 million parameters.

#### RoBERTa-large

An enhanced version of BERT, RoBERTa-large [15] has approximately 355 million parameters and includes improvements such as removal of the next sentence prediction objective, and training on expanded datasets.

#### BERTweet-large

BERTweet [16] retains the BERT architecture but is pretrained on 873 million English tweets using the RoBERTa pretraining approach, totaling approximately 355 million parameters. It has demonstrated superior performance compared to previous smaller language models in various tweet-based NLP tasks.

### Experiments

We employed a stratified sampling strategy to split the annotated dataset of 4,216 tweets into training, development, and test sets. For flu vaccine classification, we have 1,977 tweets for training, 270 for development, and 562 for testing. For flu test, we have 990 tweets for training, 135 for development, and 282 for testing.

We selected the Llama-3-70B-Instruct model for our prompting experiments, utilizing the vllm library^2^ for efficient inference. We configured the generation parameters as follows, with other parameters kept at their default values: temperature=0.2, top_*p*_=0.2. Initial prompts were drafted and subsequently refined with GPT-4o, specifically targeting task instructions and reasoning steps. Few-shot examples were chosen from the training dataset, with prompts further tuned using the development sets. We explored several prompting strategies:

#### Few-shot with CoT

The complete prompting strategy as illustrated in the Approach section.

#### Few-shot

We removed reasoning instructions from task instructions and CoT steps from examples.

#### Zero-shot with CoT

We maintained the task instruction from our complete strategy but followed a zero-shot CoT approach by adding “let’s think step by step” after each query example, as detailed in [17].

#### Zero-shot

We removed reasoning instructions from task instructions and did not include any few-shot examples.

For the baseline experiments, we employed the same classification setups used in the LLM experiments. Specifically, flu vaccine classification involved directly predicting one of the five flu vaccine labels, while flu test classification consisted of a two-step prediction process: first predicting the flu test outcome and subsequently determining the flu season. We tuned key parameters on the development set, while keep other parameters as defaults: sequence length = 256, batch size = 32, epoch size = 10, learning rate = 5e-5.

We executed all experiments on two NVIDIA A100 GPUs, each equipped with 80 GB of VRAM.

### Evaluation Metrics

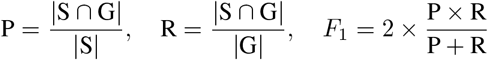

We adopted the standard evaluation metrics for multi-class classification task, Precision (P), Recall (R), and the F_1_ scores. P is defined as the ratio of correctly predicted instances (|*S* ∩ *G*|) to the total predicted instances (|*S*|), while R is the ratio of correctly predicted instances to the total ground-truth instances (|*G*|). The F_1_ score, a harmonic mean of Precision and Recall, balances the trade-off between these two metrics. In our experiments, we evaluated performance across three categories: vaccination status and test outcome of the current flu season, and the overall labels. For the evaluation of overall labels, we adopted the micro-average approach, which computes metrics by averaging over all instances.

## RESULTS

Table 4 demonstrates that Few-shot + CoT prompting achieves the highest performance compared to baseline models and other prompting methods across both flu vaccination status and flu test outcome classification tasks. Specifically, Few-shot + CoT achieves F_1_ scores of 87.35 for the *Currently-Vaccinated* and 92.14 for the *Currently-Unvaccinated*, respectively, outperforming the best baseline model (RoBERTa), which achieves 86.09 and 91.70, respectively. In flu test classification, Few-shot + CoT demonstrates an even more substantial improvement, achieving 88.89 and 89.36 in *Currently-Positive* and *Currently-Negative*, outperforming RoBERTa’s 77.27 and 81.82 F_1_ scores.

**Table 4:** F_1_ scores for baseline models and various prompting strategies in identifying flu vaccination status and flu test outcomes. The best-performing prompting strategy for each category is highlighted in bold.

|  |  | Flu Vaccination |  |  | Flu Test |  |  |
| --- | --- | --- | --- | --- | --- | --- | --- |
|  |  | Cur-Vac | Cur-Unvac | Overall | Cur-Pos | Cur-Neg | Overall |
| **Baseline** | BERT | 82.76 | 89.90 | 83.8 | 64.71 | 73.47 | 90.07 |
|  | RoBERTa | 86.09 | 91.7 | 84.48 | 77.27 | 81.82 | 91.84 |
|  | BERTweet | 84.62 | 90.46 | 84.36 | 55.17 | 75.56 | 88.56 |
| **LLM** | Zero-shot | 75.34 | 83.71 | 75.09 | 82.05 | 69.7 | 87.59 |
|  | Zero-shot + CoT | 82.21 | 84.78 | 78.83 | 81.08 | 71.70 | 90.43 |
|  | Few-shot | 80.43 | 87.63 | 80.78 | 83.72 | 86.96 | 93.26 |
|  | Few-shot + CoT | **87.35** | **92.14** | **84.88** | **88.89** | **89.36** | **95.03** |

Comparing baseline models to our best prompting strategy, we observe a discrepancy in their performances across the two classification tasks. While baseline models perform relatively well in identifying flu vaccine status, they struggle considerably with flu test classification. This discrepancy likely arises from the reliance of baseline models on the labeled training data, which for flu vaccine classification is approximately twice the size of the dataset available for flu test classification. In contrast, LLM-based prompting strategies require only a small set of labeled examples to demonstrate the task within the prompt.

Evaluating the effectiveness of different prompting strategies, we can see that incorporating CoT prompting consistently enhances performances for both Zero-shot and Few-shot prompting. For instance, adding explicit reasoning instructions (e.g., prompting the model with “let’s think step by step”) to Zero-shot prompting improves overall F_1_ scores from 75.09 to 78.83 for flu vaccination classification, and from 87.59 to 90.43 for flu test classification. Furthermore, employing a Few-shot strategy with example-driven prompting notably boosts performance compared against Zero-shot prompting, underscoring the importance of in-context examples for guiding LLM predictions. And the same effect exists when including CoT prompting. Importantly, combining Few-shot prompting with CoT reasoning results in an even more pronounced improvement. For instance, in flu vaccination classification, adding few-shot examples increases the F_1_ score by 7.36 points for the *Currently-Unvaccinated* label when CoT prompting is used, compared to only 3.92 points without CoT prompting.

## DISCUSSION

In the flu test classification task, we adopted a two-step prompting strategy, which requires the LLM to make inferences on each tweet twice. To validate the two-step process, we conducted an additional experiment combining the prompts from steps one and two into a single prompt, and instructed the LLM to directly classify each tweet into one of the five flu test labels. The single-step prompting approach achieved an F_1_ of 82.35 for the *Currently-Positive* and 84.0 for the *Currently-Negative*, representing a performance decrease of approximately 6 percentage points compared to the two-step prompting strategy. This degradation in performance can be attributed to known limitations in LLMs when handling long reasoning prompts [18, 19]. To understand what caused the degradation, we further evaluated the performances of single-step prompting in separately predicting flu test outcomes (without considering flu season) and whether the positive/negative flu test occurs in the current flu season. As shown in Table 5, the single-step approach exhibits lower performance across all three evaluation categories, with particularly notable shortcomings in accurately identifying the flu season. Since the two-step prompting needs make inference over each input example twice, we also compared the inference time (clock time) between the two-step and single-step methods. We found that the two-step prompting method required only slightly longer processing time, taking 2m 56s for 282 examples, compared to 2m 43s for the single-step method.

**Table 5:** Performance comparison between single-step and two-step classification approaches for flu test outcome prediction (positive or negative) and flu season identification.

|  |  | Positive | Negative | Current-Flu-Season |
| --- | --- | --- | --- | --- |
| Few-shot + CoT | Single-step | 88.46 | 92.86 | 85.71 |
|  | Two-step | 92.31 | 97.3 | 91.57 |

### Error Analysis

We conducted error analysis on the flu vaccination and flu test development sets to understand what errors our best prompting strategy (hereafter referred to as the model) is making. For vaccination status, the major errors come from the confusion between a user’s past behavior and their current stance. Many tweets mention having taken (or refused) a flu vaccine in previous seasons but do not explicitly state anything about the flu season or user’s action or intention for the current season. For instance, the tweet “Took one flu shot but was never sick before and sick after the shot which I never took again. Never been sick since.” was annotated as *Currently-Unvaccinated* but predicted as *Previously-Vaccinated*, although during the reasoning steps, the model explicitly implies “a habitual pattern”. Another ambiguous example is the tweet “That’s true. I did have some soreness, but it was similar to when I got the flu shot”, which was annotated with *Currently-Vaccinated* label, while the model assigned *Previously-Vaccinated* due to no clear information of the flu season.

For flu test classification, the most common errors are caused by the confusion of the temporal context. Specifically, the model often incorrectly predicted the timing of flu tests due to insufficient temporal cues. Tweets intended to indicate a current flu test status *Currently-Positive* are often mistakenly predicted as *Previously-Positive* or *Other*. For instance, the tweet “Had THAT with a negative flu test. All symptoms except no testing available” is predicted as *Other* due to insufficient temporal information about the flu test. We also observed a few annotation errors resulting from ambiguous temporal information. For instance, for the tweet “We’re pretty sure we had it in november to late December. Never been so sick, multiple visits to urgent care and the ER resulted in negative flu tests” with posting date on 2020-12-02, the model correctly interpret that “late December” (since the posting date is early December), predicting *Previously-Positive*, whereas the annotator incorrectly labeled it as *Currently-Positive*.

To estimate the real-world applicability of our approach, we evaluated its performance on the complete dataset from the 2019–2020 flu season. We stratified sampled 403 flu test tweets and 470 flu vaccine tweets based on the predicted labels for human validation. Our results indicate approximately a 1% mismatch rate between model predictions and human validation for flu test classification and a 5% mismatch rate for flu vaccination classification. Interestingly, both validation scores are better than the overall scores on our previously annotated dataset. This improvement is somewhat surprising but reasonable, given that the 2019–2020 dataset does not contain confounding or noisy information related to COVID-19 vaccination.

### Limitations and Future Work

While our few-shot CoT prompting with the LLaMA-3-70B-Instruct model achieves excellent performance, several limitations warrant further study.

First, the LLM used in this research is subject to limitations inherent to its training and architecture. Future work could explore more advanced LLMs with enhanced capabilities for processing social media language.

Second, our study relied on manually crafted prompts and a small set of few-shot examples for each label. This approach, while effective, may not always capture the optimal prompt. Future work could incorporate automatic prompt optimization techniques that are most likely to result in accurate model outputs [20, 21].

Last, our dataset is derived from historical tweets from the 2019–2020 and 2020–2021 flu seasons, which may not fully capture evolving linguistic patterns and user behaviors in social media. As a next step, we plan to apply our methodology to newly collected tweets from the 2024–2025 flu season. This will allow us to assess model robustness across different timeframes and validate its effectiveness in real-time influenza VE estimation.

### Conclusion

This study demonstrates the feasibility of using LLMs with ICL and CoT prompting to effectively extract valuable public health insights from social media data. By accurately identifying influenza test results and vaccination statuses, our approach enables real-time estimation of influenza vaccine effectiveness. Our findings show that CoT-enhanced few-shot prompting outperforms both traditional fine-tuned models and alternative LLM prompting strategies, particularly in flu test classification when temporal context is critical.

## Data Availability

All data produced will be publicly available once the paper is published

## Acknowledgments

This research was supported by the National Library of Medicine of the National Institutes of Health under Award Number 1R21LM014467-01. The content is solely the responsibility of the authors and does not necessarily represent the official views of the National Institutes of Health.

## Footnotes

2 vllm-0.6.3: https://github.com/vllm-project/vllm

